# Industry payments to anesthesiologists in the United States between 2014 and 2022

**DOI:** 10.1101/2023.07.24.23293096

**Authors:** Anju Murayama

## Abstract

**Background:** Financial relationships between physicians and the healthcare industry could be beneficial to improve patient care, but could lead to conflicts of interest. However, there was no study specifically evaluating the extent of financial relationships between anesthesiologists and the healthcare industry in the United States.

**Methods:** Using the Open Payments Database between 2014 and 2022, this longitudinal cross-sectional study examined the size, prevalence and trends of general (non-research) payments made by the healthcare industry to all anesthesiologists in the United States.

**Results:** Over the nine-year period, 67.0% of all anesthesiologists received general payments totaling $272.0 million over nine years, while 21.0% to 35.3% of anesthesiologists received one or more general payments each year. Median annual general payments to anesthesiologists ranged from $57 to $115. The top 1%, 5%, and 10% of anesthesiologists received 73.4%, 90.3%, and 94.8% of all general payments, respectively. There were no constant yearly trends in the total amounts and per-anesthesiologist general payments between 2014 and 2019, but significant declines occurred in 2020, likely due to the COVID-19 pandemic. Pain medicine physicians received the highest median general payments of $4,426 in nine-year combined total amounts, followed by addiction medicine ($431), critical care medicine ($277), and general anesthesiology ($256).

**Conclusion:** This study reveals significant financial relationships between the healthcare industry and anesthesiologists, with a disproportionate concentration of payments among a minority of anesthesiologists. While no clear trends in payments were evident before the pandemic, there was a substantial reduction during the COVID-19 outbreak.

## Main body of the manuscript

### Introduction

While certain interactions between physicians and the healthcare industry, such as sharing experiences and collaborating on research, can benefit patients through drug and device development and improved care, they may also create conflicts of interest among physicians.[1] To enhance transparency in these relationships, the United States (US) enacted the Physician Payments Sunshine Act in 2010. This act requires pharmaceutical and medical device manufacturers to disclose their financial transactions with physicians on a federal online database, the Open Payments Database, since 2014.[2] Utilizing this database, numerous previous studies have demonstrated widespread financial relationships between physicians and the healthcare industry across various specialties other than anesthesiology.[2-11]

Anesthesiologists are responsible for a considerable proportion of opioid prescriptions, accounting for 8.9% of all opioid-related prescriptions in the US, with primary care physicians and nurse practitioners prescribing the majority of opioids.[12-14] Prior research has revealed that non-research payments to physicians from opioid manufacturers were significantly associated with increased opioid prescriptions at the individual physician level[13,15-18] and higher opioid overdose deaths at the county level.[19] However, the extent of financial relationships between anesthesiologists and the healthcare industry in the US has not been well-documented. This study aims to assess the size and prevalence of industry payments to anesthesiologists.

## Methods

### Study design, participants, and data collection

This longitudinal cross-sectional study examines the extent and trends of non-research payments to all anesthesiologists in the US using the Open Payments Database. Following the Physician Payments Sunshine Act, all pharmaceutical and medical device companies are required to disclose financial transfers exceeding $100 annually or $10 per payment to physicians, nurse practitioners (since 2021), and teaching hospitals. The reported payments have been disclosed on the Open Payments Database since August 2013, but full-year payment data have been available since 2014.

All anesthesiologists in the US were identified as physicians whose primary specialty was anesthesiology and subspecialties, and their profile data were extracted from the Centers for Medicare and Medicaid Services National Plan and Provider Enumeration System database. All general payments to anesthesiologists were extracted from the Open Payments Database from 2014 to 2022, matching their National Provider Identifier number, as previously noted.[3,9,20,21]

### Data analyses

Descriptive analysis was conducted on the extracted payment data. For anesthesiologists who received payments, per-anesthesiologist payments were calculated. Additionally, yearly trends in the number of anesthesiologists receiving payments and per-anesthesiologist payments were examined using population-averaged generalized estimating equations (GEE) with robust adjustment at the individual physician level. The trend in total amounts of annual payments was examined with a linear regression model, adjusting for the impact of the COVID-19 pandemic using interrupted time series (ITS) analysis.[4,21-23] The annual number of anesthesiologists receiving payments and per-anesthesiologist payments were examined using a modified log-linked GEE model with Poisson distribution and negative binomial GEE model, respectively.[3,9,20] General payments for royalties, ownership interests, acquisitions, debt forgiveness, and long term medical supply or device loan were excluded from the trend analysis, as only small number of anesthesiologists received substantial amounts (royalties and ownership interest) or the payment categories were newly introduced since 2021 data (acquisitions, debt forgiveness, and long-term medical supply or device loan). As subgroup analyses, the study performed the GEE trend analyses excluding anesthesiologists who were deactivated and/or newly activated after 2014, to exclude new and retired physicians from the study sample. Inflation in US dollars was adjusted to 2022-dollar values in all analyses using the U.S. Bureau of Labor Statistics Consumer Price Index calculator.[3,23]

Additionally, the payments data were analyzed by payment categories, anesthesiologist subspecialties, and companies making payments. Differences in per-anesthesiologist payment and likelihood to receive payments by subspecialty were evaluated with a chi-square test and a Kruskal Wallis test. All data extraction and statistical analysis were performed using Python 3.9.12 (Python Software Foundation, Beaverton, OR, USA), Microsoft Excel, version 16.0 (Microsoft Corp., Redmond, WA, USA), and Stata version 17.0 (StataCorp, College Station, TX, USA).

### Ethical clearance

Given that the publicly available data set per the Sunshine Act reporting guidelines, Institutional board review and approval were not required for this study, as it was designed as a non-human subjects study of freely available public data.

## Results

Out of 61,305 anesthesiologists registered in the NPPES database, 67.0% (41,041) received at least one general payment from 1373 companies between 2014 and 2022, totaling $271,950,702 in inflation-adjusted amounts. Median nine-year combined total amounts of general payments per anesthesiologist were $291 (interquartile range [IQR]: $108–$974), while the mean was $6,626 (standard deviation: $66,792). The top 1%, 5%, and 10% of anesthesiologists received 73.4%, 90.3%, and 94.8% of all general payments over the nine years, respectively (Figure 1).

**Figure 1.**
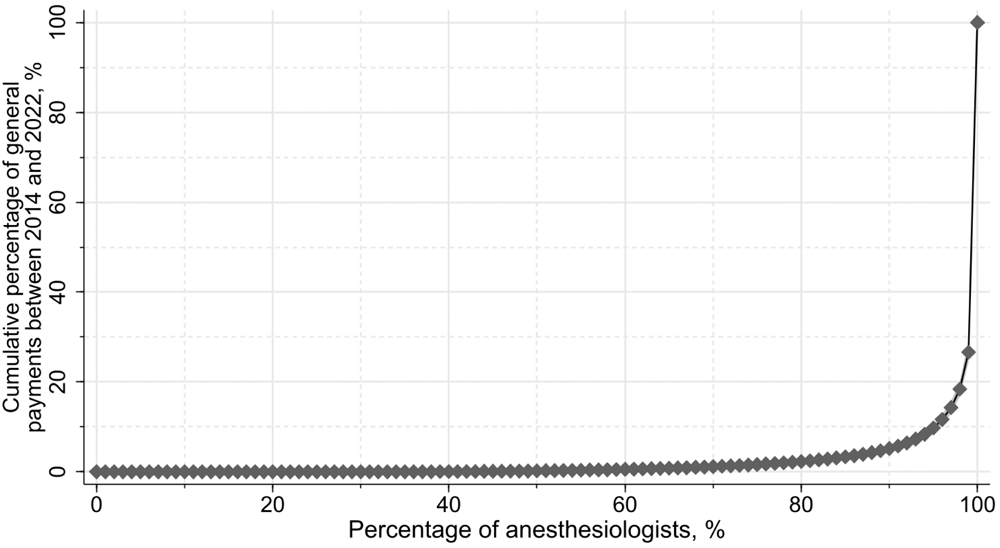
Concentration of general payments to anesthesiologists between 2014 and 2022.

### Payment trends between 2014 and 2022

Annual trends showed that 21.0% to 35.3% of anesthesiologists received one or more general payments each year between 2014 and 2022. The total amounts of general payments decreased from $37 million in 2015 to $33 million in 2019 and further to $21.7 million in 2020 during the COVID-19 pandemic. Median annual general payments to anesthesiologists ranged from $57 to $115 (Table 1).

**Table 1.** General payments to anesthesiologists between 2014 and 2022.

| Variables | Payment year |  |  |  |  |  |  |  |  | Relative annual average percentage change (95% confidence interval), %<sup>b</sup> |  |  |
| --- | --- | --- | --- | --- | --- | --- | --- | --- | --- | --- | --- | --- |
|  | 2014 | 2015 | 2016 | 2017 | 2018 | 2019 | 2020 | 2021 | 2022 | 2014-2019 | 2014-2019 vs 2020-2022 | 2020-2022 |
| Total payment amounts, \$ | 30,201,179 | 36,998,756 | 35,662,138 | 33,336,258 | 33,964,108 | 32,988,544 | 21,705,999 | 23,302,673 | 23,791,047 | 0.8 (-2.6 to 4.3) | -72.4 (-84.4 to -60.4)* | 9.9 (5.1 – 14.7)* |
| Number of physicians with payments, n (%) | 17,061 (27.8) | 18,177 (29.7) | 21,642 (35.3) | 19,466 (31.8) | 19,215 (31.3) | 19,245 (31.4) | 12,887 (21.0) | 16,277 (26.6) | 17,709 (28.9) | 1.8 (1.5 – 2.0)* | -43.2 (-44.2 to -42.1)* | 14.7 (13.8 – 15.6)* |
| Payments per physician, \$<sup>c</sup> | | | | | | | | | | | | |
| Median (IQR) | 112 (33–280) | 115 (35–306) | 115 (34–282) | 104 (32–270) | 99 (30–255) | 109 (30–255) | 57 (22–197) | 74 (25–213) | 68 (24–211) | 0.7 (-2.4 to 3.9)<sup>c</sup> | -59.5 (-65.2 to -52.9)*<sup>c</sup> | 17.2 (9.7 – 25.1)*<sup>c</sup> |
| Average (SD) | 1,770 (16,773) | 2,035 (26,148) | 1,648 (13,435) | 1,713 (14,203) | 1,768 (15,892) | 1,714 (19,874) | 1,684 (32,580) | 1,432 (14,750) | 1,343 (17,421) |  |  |  |

Legend: <sup>a</sup> Per-physician payment was calculated among physicians receiving payments. <sup>b</sup> General payments for royalties, ownership interests, acquisitions, debt forgiveness, and long-term medical supply or device loan were excluded from the trend analysis, as only small number of anesthesiologists received substantial amounts (royalties and ownership interest) or the payment categories were newly introduced since 2021 data (acquisitions, debt forgiveness, and long-term medical supply or device loan). <sup>c</sup> These values were estimated relative annual average percentage change among all anesthesiologists registered in the NPPES database including those who did not receive any payments.
\*p<0.001. Abbreviations: 95% confidence interval (95% CI), interquartile range (IQR), SD (standard deviation)

The GEE model indicated no constant trends in the amounts, counts, and per-anesthesiologist general payments between 2014 and 2019 (Table 1). However, the number of anesthesiologists with general payments increased by 1.8% (95% confidence interval [95% CI]: 1.5–2.0, p<0.001) each year. Total annual amounts of general payments significantly decreased by 72.4% (95% CI: -84.4% to -60.4%, p<0.001) in 2020 compared to those between 2014 and 2019. Similarly, both the number of anesthesiologists receiving general payments and per-anesthesiologist payment amounts significantly decreased by 43.2% (95% CI: -44.2% to -42.1%, p<0.001) and 59.5% (95% CI: -65.2% to -52.9%, p<0.001) in 2020, respectively. However, there were significant recovery trends in all variables between 2020 and 2022.

Subgroup analyses excluding newly activated anesthesiologists showed similar trends in the total annual amounts of payments and per-anesthesiologist payment amounts (Supplemental Material 1). The number of anesthesiologists receiving general payments slightly increased by 0.5% (95% CI: 0.3%–0.8%, p<0.001), indicating that the increasing trend observed in the previous GEE model was primarily due to new anesthesiologists receiving general payments.

### Payments by category and company

In payment amount, speaking compensation not related to continuing medical education (CME) program accounted for the largest proportion (29.3%) of all general payments, followed by consulting payments (20.9%). Food and beverage payments made up 15.5% of all general payments but were received by 66.2% of anesthesiologists.

In payment amounts, speaking compensation not related to continuing medical education (CME) program is the largest payment categories occupying 29.3% ($79.7 million) of all general payments, followed by consulting payments (20.9%, $56.8 million) (Supplemental Material 2). Food and beverage payments accounted for 15.5% ($42.2 million) of all general payments, but were received by 66.2% of all anesthesiologists and the number of food and beverage payments occupied 87.5% (1,183,671) of general payments. Only 3.2% (1,944) and 4.9% (2,988) of anesthesiologists received the speaking compensations not related to CME program and consulting payments, respectively. The highest per-payment values were $111,162 in acquisition payments, $31,687 in royalty or license payments, $24,662 in ownership payments. Per-payment value were $2,976 in consulting payments and $2,772 in speaking compensations for non-CME program, while it was $36 in food and beverage payments.

Among 1373 companies, the top 10 and 30 companies accounted for 41.9% ($113.9 million) and 67.8% ($184.5 million) of all general payments, respectively (Supplemental Material 3). Pacira Pharmaceuticals made the largest general payments ($19.8 million), followed by Medtronic USA ($15.1 million), Boston Scientific ($13.7 million), Halyard Health ($11.2 million), and Abbott Laboratories ($11.0 million).

### Payments by subspecialty

The proportion of anesthesiologists receiving general payments over the nine-year period was highest in pain medicine (89.5%), followed by addiction medicine (69.5%) and general anesthesiology (65.7%) (Figure 2). Pain medicine physicians received the highest median general payments of $4,426 (IQR: $1,077–$12,881), followed by addiction medicine ($431 [IQR: $177–$2,030]), critical care medicine ($277 [IQR: $113–$1,081]), and general anesthesiology ($256 [IQR: $98–$717]) (Figure 3). There were significant differences by subspecialty in the proportion of anesthesiologists receiving one or more general payments over the nine-years (p<0.001 in chi-square test) and per-anesthesiologist general payments (p<0.001 in Kruskal Wallis test). While pain medicine physicians were 6.8% (4,164 out of 61,305) of all anesthesiologists included in this study, they received 38.1% ($103.6 million) of all general payments from the healthcare industry.

**Figure 2.**
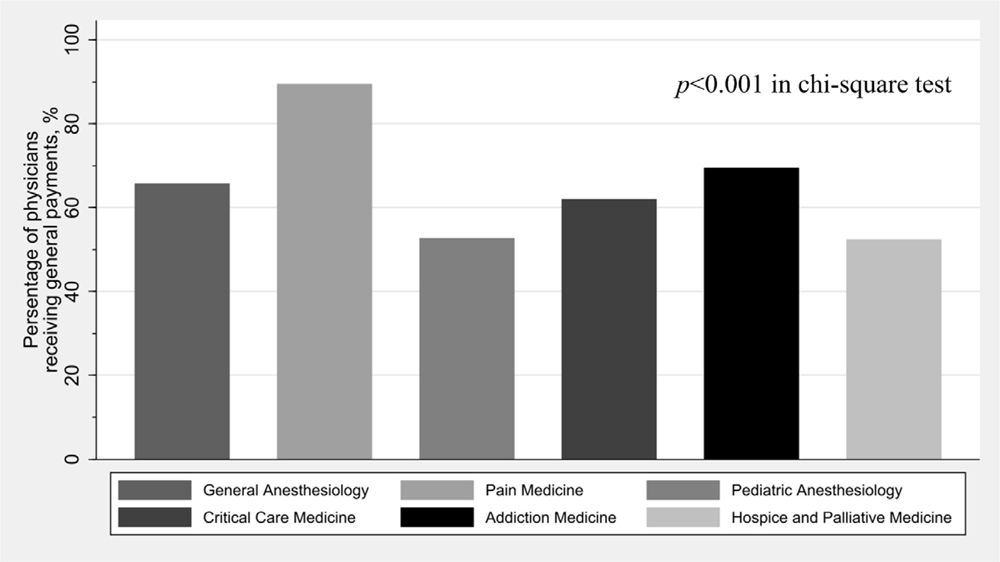
Proportions of anesthesiologists receiving at least one general payment between 2014 and 2022 by subspecialty.

**Figure 3.**
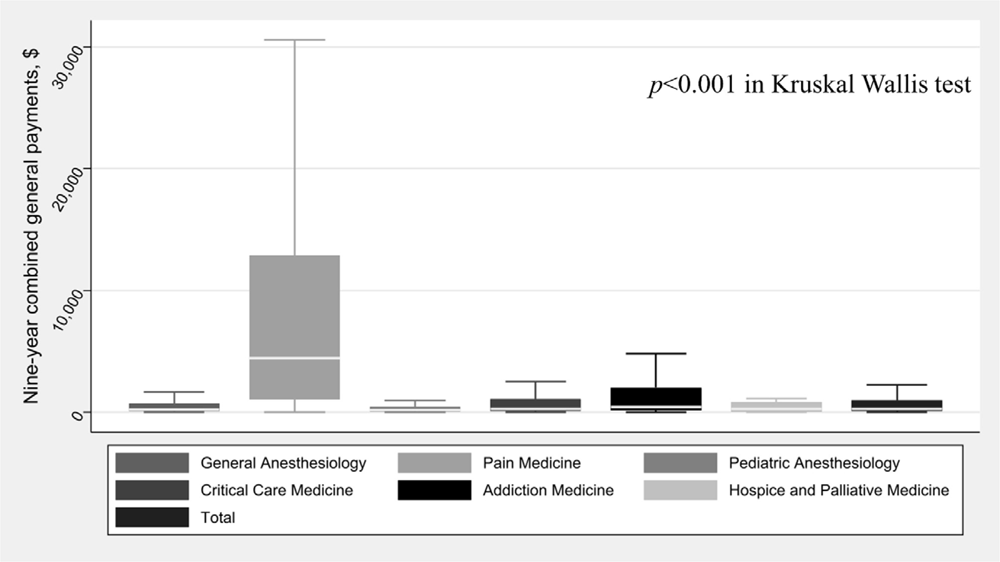
Median general payments per anesthesiologist among anesthesiologists who received general payments between 2014 and 2 by subspecialty.

Regarding payment trends by subspecialty, only pain medicine exceeded the majority of anesthesiologists receiving general payments each year (Supplemental Material 4). Median per-anesthesiologist general payments were the highest in pain medicine ($326–$732), followed by addiction medicine ($71–$196) and critical care medicine ($69–$139).

## Discussion

This study is the first analysis to assess the magnitude and extent of general payments provided to all anesthesiologists in the United States. The findings demonstrate that more than $271 million in general payments were made to 67.0% of all anesthesiologists over the nine-year period, with only a minority of anesthesiologists receiving payments each year. Moreover, a large proportion of the general payments were received by a small number of anesthesiologists. The study also reveals differences in general payment patterns within anesthesiology subspecialties, with pain medicine physicians receiving the highest median general payments, primarily from pharmaceutical companies marketing opioids. Pain medicine physicians received more than 17 times higher general payments in median amounts than general anesthesiologists. Although there were no continuing trends in payments between 2014 and 2019, both the number of anesthesiologists receiving general payments and per-anesthesiologist general payments significantly decreased in 2020. Compared to the findings from previous studies, this study would provide important insight into the transparency of the financial relationship between anesthesiologists and the healthcare industry.

First, this study found that the median annual general payments to anesthesiologists ranged from $57 to $115 each year. Compared to previous studies, this study indicates that anesthesiologists received fewer general payments from the healthcare industry than other medical specialists in the US. For instance, Marshall and her colleagues reported that median general payments per physician were $186 in family medicine physicians, $251 in general surgeons, $453 in colorectal surgeons, and $860 in thoracic surgeons in 2015.[2] Another studies based on the same study design among internal medicine subspecialties reported that median annual general payments were $197 to $220 in infectious disease specialists[6], $379 to $440 in pulmonologists[3], $395–$590 in allergy and clinical immunology[4], $632 in medical oncology[24], $730 to $812 in rheumatologists[21], $725 to $847 in cardiologists.[22] Additionally, given mean annual wage for anesthesiologists was $302,970 according to 2022 estimates from the U.S. Bureau of Labor Statistics,[25] the majority of anesthesiologists have not received large general payments compared to their annual salary as an anesthesiologist.

An intriguing observation from this study is the significant variation in general payments among anesthesiology subspecialties. Notably, pain medicine physicians received substantially higher median general payments compared to other anesthesiologists and even certain specialists from different medical fields. Median nine-year payment amounts to pain medicine physicians were 17.3 times larger than those to general anesthesiologists ($4,426 vs $256). Additionally, compared to other specialists, median annual general payments to pain medicine physicians ($326–$732) were as same as or higher than several surgery and internal medicine specialists such as general surgeons,[2] pulmonologists,[3] allergists,[4] and medical oncologists.[2,24] These large payments to pain medicine physicians were made by pharmaceutical companies marketing opioids in the United States, such as INSYS Therapeutics, Depomed, Purdue Pharma, and Pfizer. Intense marketing activities by opioid manufacturers to physicians and healthcare community have been well documented since the early 2000s.[15-19,26] Hadland et al. reported that pain medicine physicians received the third largest general payments related to opioid of all specialists between 2013 and 2015.[18] This finding warrants further investigation and raises questions about the potential influence of these financial relationships on pain medicine practice and decision-making. Given the well-documented association between opioid-related payments and increased opioid prescriptions, healthcare costs,[17,19,26] and opioid overdose deaths,[19] it is crucial for pain medicine physicians to be cautious and transparent in their financial interactions with opioid manufacturers. While this study does not establish any undue influence of industry payments on anesthesiologists, it underscores the importance of vigilance and adherence to conflicts of interest guidelines to maintain the integrity of clinical, education, and research practices.[1,27-29] Further research is warranted to explore the implications of these financial interactions on clinical decision-making, healthcare practices, and patient outcomes.

Furthermore, only the top 1% and 10% of anesthesiologists received nearly three fourths and 95% of all general payments, respectively. This finding is consistent with many previous studies in this research field.[3,5,8–11,21–23,30-34] Feng et al. reported that the top 10% of dermatologists received more than 90% of all general payments from the healthcare industry in 2014.[8] The top 10% of pulmonologists received 83.7% of all general payments between 2013 and 2021.[3] As shown in previous studies, the healthcare industry made the large amounts of general payments to the small fraction of physicians who are in leading positions such as clinical guideline authors,[7,32,35-37] journal editors,[32,37-40] society board members,[32,41] and principal investigators of industry-sponsored clinical trials.[28,42] These influential physicians are called key opinion leaders.[42,43] Although some studies positively stated the role of key opinion leaders who are supported by the healthcare industry when they advocate as an independent expert for better, cheaper and safer products on behalf of patients and healthcare communities,[33] most of previous cases have not shown such positive effects. Instead, financial conflicts of interest between healthcare industry and key opinion leaders have questioned the influence of experts on recommendations and practices.[27,44-47] Current global standards for managing conflicts of interest require these influential physicians such as clinical guideline authors and journal editors to minimize their financial interactions with the healthcare industry and at least to be transparent in their relationships with the healthcare industry. Nevertheless, Wyssa et al. found that there was no disclosure of authors’ conflicts of interest in 36% of clinical guidelines published in ten high-impact anesthesiology academic journals between 2008 and 2018.[48] Additionally, some studies in other specialties found that there were large amounts of undeclared and under-declared financial relationships between the healthcare industry and influential physicians.[49] Further transparency in financial relationships between the healthcare industry and the influential physicians is needed. Future studies should evaluate the extent and accuracy of disclosed financial relationships between the healthcare industry and influential anesthesiologists.

Another notable aspect of this study is its evaluation of trends in general payments over time. Despite the implementation of the Open Payments Database to increase transparency in physicians-industry relationships, some experts predicted that increased transparency and increased public pressure and scrutiny would lead to a decrease in physicians-industry financial relationships for non-research purposes.[50] This study found limited changes in the general payments patterns between 2014 and 2019. This observation would raise questions about the effectiveness of the Open Payments Database in altering anesthesiologists-industry financial relationships. However, the COVID-19 pandemic had a profound impact on general payments in 2020, with a substantial decrease in both the amounts and the number of anesthesiologists receiving payments. This finding is consistent with previous studies in other specialties and overall.[6,21,22,30,31,51] Inoue et al. found that the amounts of general payments decreased by more than 44% overall in 2020 compared to those in 2019.[51] Santamaria et al. reported that general payments to surgeons decreased by about 50% in 2020.[30] This decline in general payments to anesthesiologists can be attributed to the pandemic-related restrictions and measures that disrupted traditional marketing strategies and in-person interactions. Interestingly, the industry seemed to adapt quickly to the changing landscape, as evidenced by the significant recovery trends in payments between 2020 and 2022, while the total amount of general payments to anesthesiologists in 2022 was still about 70% of those before the pandemic ($23.8 million in 2022 vs $33.0 million in 2019). This adaptation likely involved a shift towards utilizing online platforms and virtual engagement to maintain communication and marketing efforts during the pandemic.[21,22,30] Considering the current significant downsizing of pharmaceutical representatives, the use of online platforms and virtual marketing will continue even after the pandemic.

The limitations of this study include the possibility of errors in public databases, such as the NPPES and Open Payments Database, as pointed out in previous studies.[3,21] Second, there would be unmeasured confounding factors that may impact trends in general payments to anesthesiologists.[3,9] Third, there would be differences in the number of anesthesiologists between the U.S. Centers for Medicare & Medicaid Services NPPES database and other databases such as the American Board of Anesthesiology and the Association of American Medical Colleges databases, as these are structured based on different methods and physicians self-declared their primary specialty in the NPPES database even though they did not have board-certification as an anesthesiologist.[2,20] Fourth, the amounts of general payments to anesthesiologists would be influenced by non-measured anesthesiologists’ demographic factors such as physician age, workplace type, position, and graduated school. However, these variables are not available from the NPPES and the Open Payments Database. Finally, there would be unmeasured interactions such as free drug samples not covered by the Open Payments Database and the Physician Payments Sunshine Act.

In summary, this study reveals that over $271 million in general payments were made to 67.0% of anesthesiologists between 2014 and 2022, with only a minority receiving these payments annually. The majority of payments were received by the small group of anesthesiologists. While there were no significant trends in payments from 2014 to 2019, the COVID-19 pandemic caused a notable decline in 2020. However, payments showed a remarkable recovery between 2020 and 2022. These findings emphasize the importance of transparency in financial relationships and the need to address potential conflicts of interest. Continued efforts to promote transparency are essential for maintaining trust in healthcare practices.

## Data Availability

All data produced are available online at the Open Payments Database.

https://openpaymentsdata.cms.gov/

https://npiregistry.cms.hhs.gov/search

## Acknowledgments

I would like to thank Ms. Megumi Aizawa for her dedicated support of my research. I declared no financial conflicts of interest and did not receive any financial support for this study from any entity. For proofreading parts of the presented text, I used the freely available pre-trained ChatGPT (version 3.5) model developed by OpenAI in order to check and proofread the manuscript for language, spelling and grammatical errors. I checked and edited the text for unintended plagiarism, and verified all facts and values that I used from the ChatGPT outputs before the manuscript submission.

---

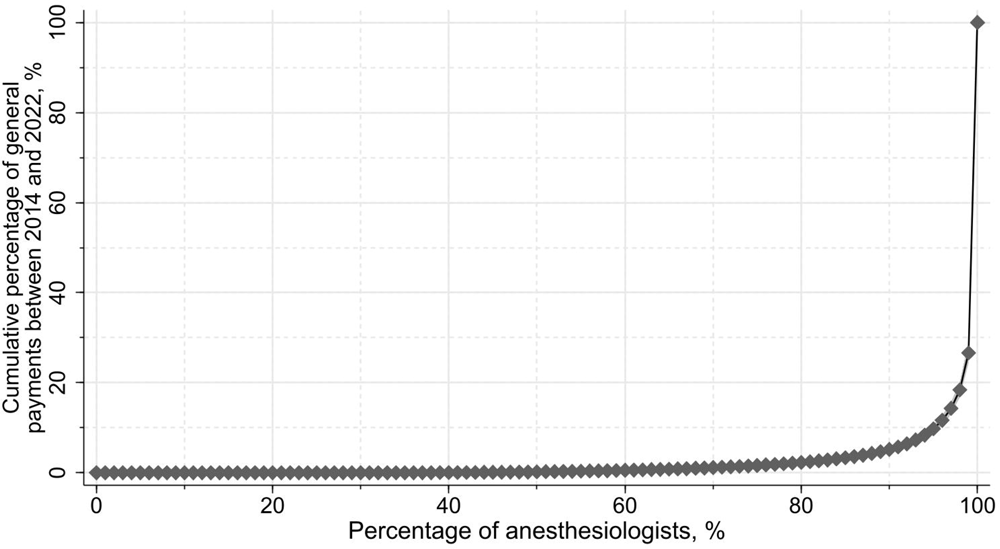

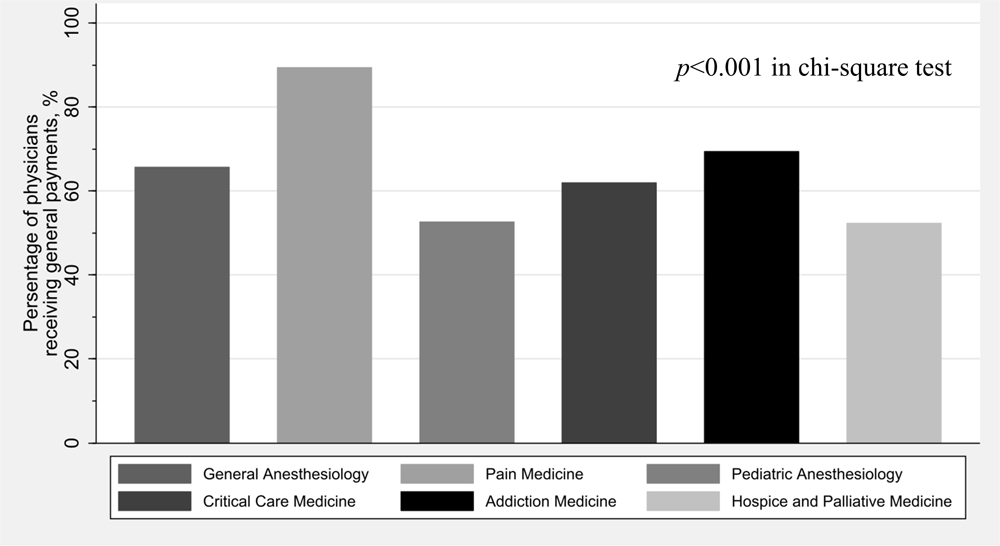

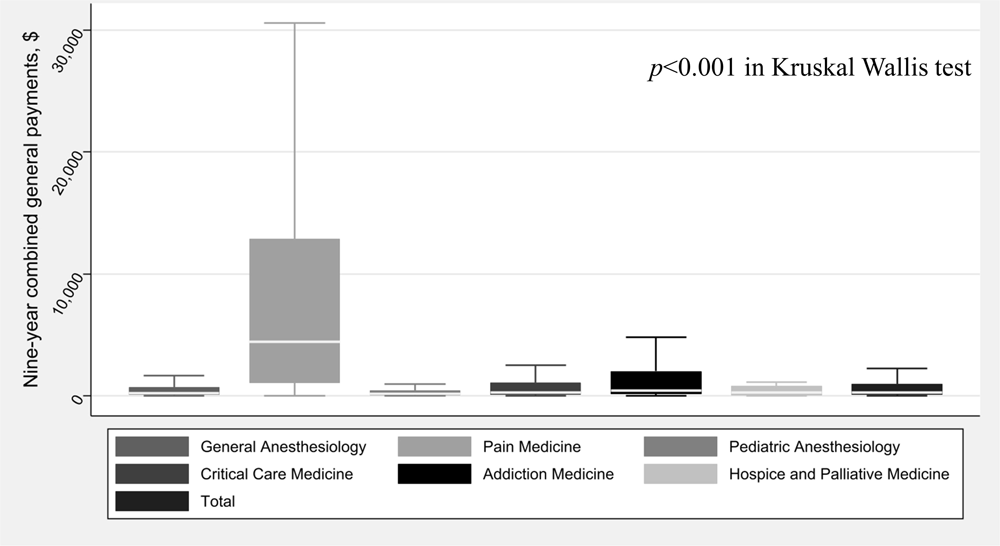

## Notes

### Competing Interest Statement

The authors have declared no competing interest.

### Funding Statement

This study did not receive any funding

