## Supplemental Materials for "Industry payments to anesthesiologists in the United States between 2014 and 2022"

Anju Murayama^1,2*^

**Affiliations:**

^1^ School of Medicine, Tohoku University, Sendai City, Miyagi, Japan

^2^ Department of Population Health Science and Policy, Icahn School of Medicine at Mount Sinai, New York City, NY, USA

Supplemental material 1. Trends in general payments to anesthesiologists between 2014 and 2022

| Variables | All anesthesiologists | | | Anesthesiologists who were continuously activated between 2014 and 2022 | | |
| --- | --- | --- | --- | --- | --- | --- |
|  | Relative annual average percentage change (95% confidence interval), % ^a^ | | | Relative annual average percentage change (95% confidence interval), %^a^ | | |
|  | 2014-2019 | 2014-2019 vs 2020-2022 | 2020-2022 | 2014-2019 | 2014-2019 vs 2020-2022 | 2020-2022 |
| Total payment amounts, $ | 0.8 (-2.6 to 4.3) | -72.4 (-84.4 to -60.4)** | 9.9 (5.1 – 14.7)** | -0.1 (-4.0 to 3.9) | -72.1 (-85.7 to -58.5)** | 8.8 (3.5 – 14.1)* |
| Number of physicians with payments, n (%) | 1.8 (1.5 – 2.0)** | -43.2 (-44.2 to -42.1)** | 14.7 (13.8 – 15.6)** | 0.5 (0.3 – 0.8)** | -43.8 (-44.8 to -42.7)** | 13.5 (11.6 – 13.5)** |
| Payments per physician, $^a^ |  |  |  |  |  |  |
| Median (IQR) | 0.7 (-2.4 to 3.9) | -59.5 (-65.2 to -52.9)** | 17.2 (9.7 – 25.1)** | -0.1 (-3.2 to 3.1) | -60.0 (-65.8 to -53.1)** | 15.6 (7.9 – 23.8)** |
| Average (SD) |  |  |  |  |  |  |

Legend: ^a^ General payments for royalties, ownership interests, acquisitions, debt forgiveness, and long term medical supply or device loan were excluded from the trend analysis, as only small number of anesthesiologists received substantial amounts (royalties and ownership interest) or the payment categories were newly introduced since 2021 data (acquisitions, debt forgiveness, and long-term medical supply or device loan). ^c^ These values were estimated relative annual average percentage change among all anesthesiologists registered in the NPPES database including those who did not receive any payments, not only among anesthesiologists receiving general payments. *p<0.01. **p<0.001. 95% confidence interval (95% CI).

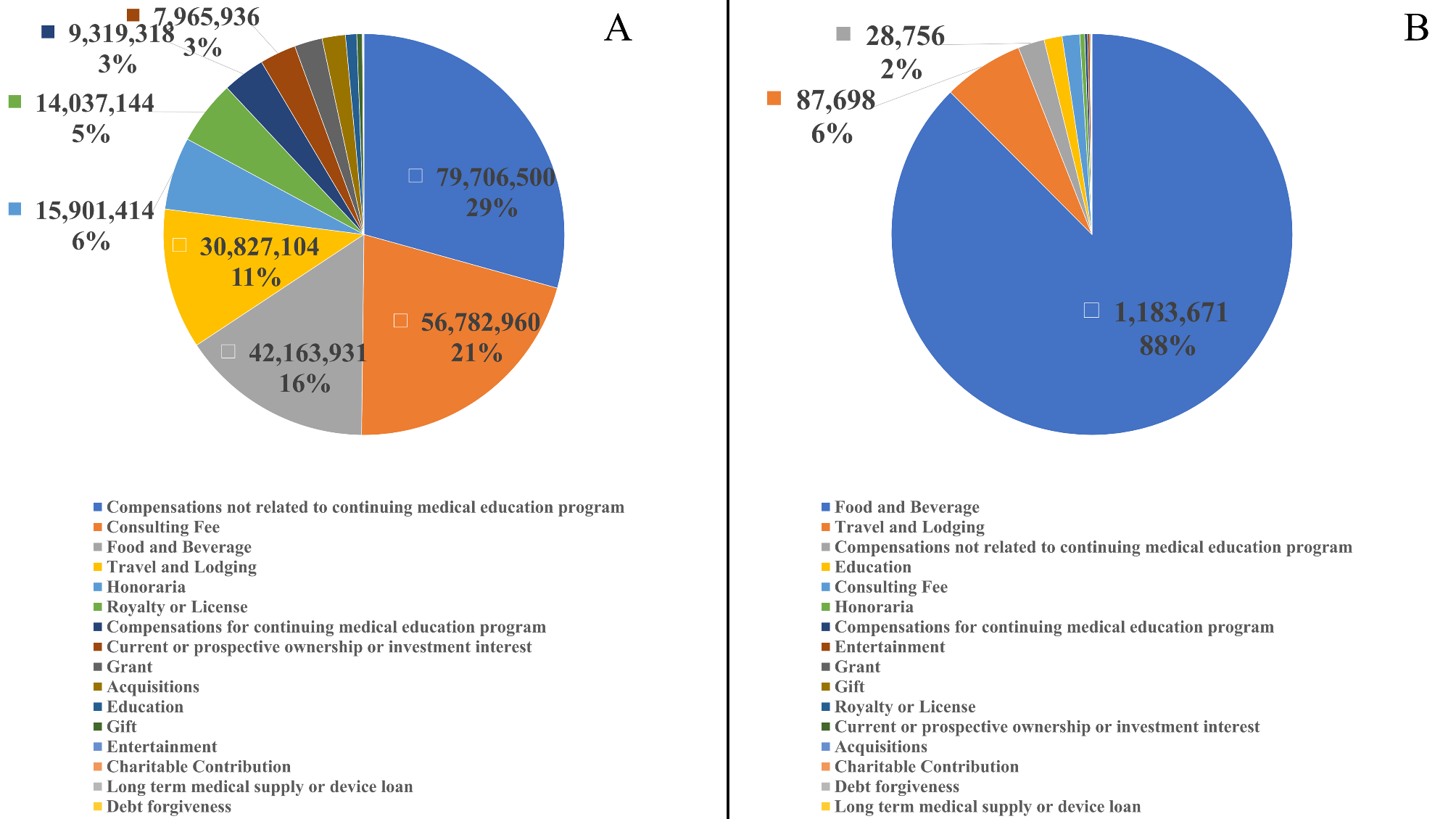

Supplemental Material 2. Total amounts and number of general payments to anesthesiologists between 2014 and 2022 by payment category

Supplemental Material 3. General payments from 30 top-paying companies between 2014 and 2022

| Ranking | Company name | Nine-year combined total amounts of general payments to anesthesiologists | |
| --- | --- | --- | --- |
|  |  | Payment amounts, US$ | Percentage of general payments to all general payments, % |
| 1 | Pacira Pharmaceuticals Incorporated | 19,773,345 | 7.3 |
| 2 | Medtronic USA, Inc. | 15,058,660 | 5.5 |
| 3 | Boston Scientific Corporation | 13,666,319 | 5.0 |
| 4 | Halyard Health, Inc. | 11,189,407 | 4.1 |
| 5 | Abbott Laboratories | 10,979,348 | 4.0 |
| 6 | Merck Sharp & Dohme Corporation | 9,263,305 | 3.4 |
| 7 | Edwards Lifesciences Corporation | 9,229,925 | 3.4 |
| 8 | Masimo Corporation | 8,603,012 | 3.2 |
| 9 | Nevro Corp. | 8,428,026 | 3.1 |
| 10 | Medtronic, Inc. | 7,705,631 | 2.8 |
| 11 | Mallinckrodt LLC | 6,283,378 | 2.3 |
| 12 | INSYS Therapeutics Inc | 5,796,607 | 2.1 |
| 13 | Depomed, Inc. | 5,711,137 | 2.1 |
| 14 | Purdue Pharma L.P. | 4,611,904 | 1.7 |
| 15 | St. Jude Medical, Inc. | 4,574,428 | 1.7 |
| 16 | Medtronic Vascular, Inc. | 4,035,604 | 1.5 |
| 17 | Pfizer Inc. | 3,784,739 | 1.4 |
| 18 | Covidien LP | 3,560,811 | 1.3 |
| 19 | Daiichi Sankyo Inc. | 3,479,612 | 1.3 |
| 20 | Baxter Healthcare | 3,443,320 | 1.3 |
| 21 | Advanced Circulatory Systems Inc. | 3,339,064 | 1.2 |
| 22 | Smith & Nephew, Inc. | 2,869,985 | 1.1 |
| 23 | CSL Behring | 2,838,490 | 1.0 |
| 24 | AstraZeneca Pharmaceuticals LP | 2,679,610 | 1.0 |
| 25 | The Medicines Company | 2,669,267 | 1.0 |
| 26 | Allergan Inc. | 2,435,235 | 0.9 |
| 27 | Collegium Pharmaceutical, Inc. | 2,242,066 | 0.8 |
| 28 | Stryker Corporation | 2,158,825 | 0.8 |
| 29 | Valeant Pharmaceuticals North America LLC | 2,048,658 | 0.8 |
| 30 | Philips Electronics North America Corporation | 2,024,003 | 0.7 |

Legend: Company names were differentiated by companies making general payments based on information disclosed in the Open Payments Database. To increase the accuracy of the companies making payments, payments from subsidiaries and affiliates were not consolidated into a single company in this study.

Supplemental Material 4. General payments to anesthesiologists by subspecialty between 2014 and 2022

| Variables | Payment year | | | | | | | | | Relative annual average percentage change (95% confidence interval), %^c^ | | |
| --- | --- | --- | --- | --- | --- | --- | --- | --- | --- | --- | --- | --- |
|  | 2014 | 2015 | 2016 | 2017 | 2018 | 2019 | 2020 | 2021 | 2022 | 2014-2019 | 2014-2019 vs 2020-2022 | 2020-2022 |
| Number of anesthesiologists receiving payments, n (%) |  |  |  |  |  |  |  |  |  |  |  |  |
| General Anesthesiology (n=54,324) | 14,265 (26.3) | 15,196 (28.0) | 18,289 (33.7) | 16,197 (29.8) | 15,960 (29.4) | 15,964 (29.4) | 10,125 (18.6) | 13,191 (24.3) | 14,480 (26.7) | 1.6 (1.3 – 1.8)*** | - 46.9 (-48.1 to -45.8)*** | 17.1 (16.0 – 18.2)*** |
| Pain Medicine (n=4164) | 2,305 (55.4) | 2,456 (59.0) | 2,688 (64.6) | 2,659 (63.9) | 2,654 (63.7) | 2,705 (65.0) | 2,439 (58.6) | 2,591 (62.2) | 2,674 (64.2) | 2.9 (2.4 – 3.4)*** | -15.3 (-17.6 to -12.9)*** | 1.7 (0.5 – 3.0)** |
| Pediatric Anesthesiology (n=1579) | 252 (16.0) | 260 (16.5) | 348 (22.0) | 298 (18.9) | 273 (17.3) | 249 (15.8) | 144 (9.1) | 233 (14.8) | 261 (16.5) | -0.3 (-2.5 to 2.1) | -57.3 (-64.7 to -48.3)*** | 32.5 (21.8 – 44.1)*** |
| Critical Care Medicine (n=1101) | 198 (18.0) | 223 (20.3) | 261 (23.7) | 270 (24.5) | 275 (25.0) | 286 (26.0) | 155 (14.1) | 234 (21.3) | 259 (23.5) | 7.1 (4.6 – 9.7)*** | -56.2 (-63.4 to -47.5)*** | 19.0 (10.5 – 28.2)*** |
| Addiction Medicine (n=95) | 36 (37.9) | 36 (37.9) | 46 (48.4) | 33 (34.7) | 44 (46.3) | 34 (35.8) | 23 (24.2) | 22 (23.2) | 28 (29.5) | 0.1 (-4.5 to 4.8) | -48.4 (-67.0 to -19.2)** | 10.8 (-9.9 to 36.2) |
| Hospice and Palliative Medicine (n=42) | 5 (11.9) | 6 (14.3) | 10 (23.8) | 9 (21.4) | 9 (21.4) | 7 (16.7) | 1 (2.4) | 6 (14.3) | 7 (16.7) | 6.9 (-8.7 to 25.2) | -88.8 (-97.8 to -43.4)** | 86.5 (0.0 – 247.7)* |
| Overall | 17,061 (27.8) | 18,177 (29.7) | 21,642 (35.3) | 19,466 (31.8) | 19,215 (31.3) | 19,245 (31.4) | 12,887 (21.0) | 16,277 (26.6) | 17,709 (28.9) | 1.8 (1.5 – 2.0)*** | -43.2 (-44.2 to -42.1)*** | 14.7 (13.8 – 15.6)*** |
| Median payments per anesthesiologist (interquartile range), $^a^ |  |  |  |  |  |  |  |  |  |  |  |  |
| General Anesthesiology | 91 (30–199) | 94 (32–220) | 97 (31–209) | 81 (28–181) | 77 (27–170) | 83 (26–174) | 41 (20–137) | 54 (22–148) | 52 (22–144) | 2.6 (-1.2 to 6.6) | -57.7 (-65.9 to -47.6)*** | 11.3 (1.2 – 22.3)* |
| Pain Medicine | 647 (176–1,814) | 638 (180–1,938) | 679 (166–2,038) | 732 (173–2,050) | 685 (174–2,162) | 592 (154–2,019) | 326 (96–1,020) | 428 (124–1,311) | 448 (132–1,419) | -2.4 (-7.7 to 3.2) | -60.5 (-68.0 to -51.2)*** | 26.4 (15.5 – 38.4)*** |
| Pediatric Anesthesiology | 76 (27–159) | 92 (22–169) | 57 (23–151) | 76 (26–151) | 71 (24–142) | 78 (25–149) | 35 (19–141) | 56 (20–153) | 34 (19–125) | 5.4 (-8.7 to 21.6) | -65.8 (-83.9 to -27.6)** | 48.5 (-3.7 to 128.8) |
| Critical Care Medicine | 125 (40–371) | 139 (54–413) | 108 (35–219) | 118 (33–326) | 115 (39–350) | 115 (30–293) | 69 (25–250) | 106 (27–245) | 110 (30–230) | 12.2 (-2.5 to 29.2) | -74.2 (-90.8 to -27.9)* | -1.7(-25.8 to 30.2) |
| Addiction Medicine | 185 (37–780) | 196 (45–720) | 128 (32–533) | 184 (58–1,080) | 146 (65–311) | 71 (41–250) | 92 (16–226) | 89 (19–175) | 77 (29–184) | -2.2 (-31.2 to 38.9) | -59.7 (-74.2 to -37.1)*** | 14.0 (-21.5 to 65.5) |
| Hospice and Palliative Medicine | 96 (69–134) | 175 (110–204) | 91 (28–126) | 207 (41–374) | 66 (19–87) | 139 (96–163) | 32 (not applicable)^b^ | 86 (38–112) | 164 (90–264) | 9.1 (-2.1 to 21.7) | -99.4 (-99.9 to -96.7)*** | 474.6 (214.2 – 950.8)*** |
| Overall | 112 (33–280) | 115 (35–306) | 115 (34–282) | 104 (32–270) | 99 (30–255) | 109 (30–255) | 57 (22–197) | 74 (25–213) | 68 (24–211) | 0.7 (-2.4 to 3.9) | -59.5 (-65.2 to -52.9)*** | 17.2 (9.7 – 25.1)*** |

Legend: ^a^ Per-anesthesiologist payment was calculated among physicians receiving payments. ^b^ Only one anesthesiologist received general payments in 2020, and interquartile range was not able to be calculated. ^c^ General payments for royalties, ownership interests, acquisitions, debt forgiveness, and long-term medical supply or device loan were excluded from the trend analysis, as only small number of anesthesiologists received substantial amounts (royalties and ownership interest) or the payment categories were newly introduced since 2021 data (acquisitions, debt forgiveness, and long-term medical supply or device loan). *p<0.05. **p<0.01. ***p<0.001.
